# Genetic study of late-onset neonatal sepsis reveals significant differences by sex

**DOI:** 10.1101/2021.08.12.21261209

**Authors:** Timothy H. Ciesielski, Xueyi Zhang, Alessandra Tacconelli, Irja Lutsar, Vincent Meiffredy de Cabre, Emmanuel Roilides, Cinzia Ciccacci, Paola Borgiani, William K. Scott, NeoMero Consortium, Scott M. Williams, Giorgio Sirugo

## Abstract

Late-Onset Neonatal Sepsis (LOS) is a rare, but life-threatening condition, involving widespread infection, immune disruption, organ dysfunction, and often death. Because exposure to pathogens is not 100% preventable, identifying susceptibility factors is critical to defining neonates at greater risk. Prior studies demonstrated that both genetics and infant sex influence susceptibility. We, therefore, performed a genome wide association study (GWAS) with 224 cases and 273 controls from six European countries to identify genetic risk factors. We tested for association with both autosomal and X-chromosome variants in the total sample and in the samples stratified by sex. In total 71 SNPs associated with neonatal sepsis at p<1×10^−4^ in at least one analysis. Most importantly, the sex stratified analyses revealed associations with multiple SNPs (28 SNPs in males and 16 in females), but none of 44 SNPs from single-sex analyses associated with sepsis in the other sex at p<0.05, and the vast majority showed significant SNP X sex interactions (17 of 28 from the male-only analyses and all 16 from the female-only study). Pathway analyses showed that NOTCH signaling is over-represented among loci identified by the analyses. Our results indicate that susceptibility to LOS is genetically sexually dimorphic, and suggest that NOTCH signaling is likely to play a role in it.

## Introduction

Late-Onset Neonatal Sepsis (LOS) is a serious infection response that develops after the third day of life.^1 2^ In this rare disorder, the neonate develops widespread infection, diffuse and ineffective immune responses, and cardiovascular instability that leads to organ failure and death in approximately 15-30% of cases.^1-3^ In high income countries this condition affects approximately 2-4 of 1000 live births^4^, but in low and middle income countries the picture is far less clear due to worse surveillance.^5 6^ There are many reasons that neonatal infections may be more common in poor resource settings; hospital hygiene is less stringent and effective treatments are less available than in upper-middle and high income countries.^5 6^ LOS can only occur when infants are exposed to infectious agents in the first few days of life, and a variety of procedures and clinical practices can prevent these contacts. ^5 7^ However as virulent pathogens are not completely avoidable, it is critical to identify host susceptibility factors, learn more about the pathophysiology, and develop interventions that can help the neonates at greatest risk.

Neonatal sepsis is a phenotypically complex and heterogeneous disorder but there is evidence that both sex^8-12^ and genetics^13^ modulate risk. Males appear to be more susceptible to LOS as immune function and infection susceptibility differs between the sexes at all ages.^8-12^ Thus, it is possible that at least some of the etiologic processes and genetic risk factors differ by sex. Therefore, we performed a genome wide association study (GWAS) to identity variants that associate with neonatal sepsis in sex-combined and sex-stratified analyses. This approach is critical in the context of traits with sex differences ^8 10 14-16^, and is in line with recent consensus statements and published analytic guidelines^16-20^ that recommend considering the sexes independently in biomedical research. Failing to stratify by sex might impair the recognition of context dependent risk factors because the biology may differ.

Overall, we designed our analyses to embrace the etiologic heterogeneity of neonatal sepsis with the goal of better understanding pathophysiology to develop effective new interventions.

## Methods

### Data Source

Neonates with LOS were identified in 21 hospitals across Europe as previously described as part of the NeoMero studies.^1 21^ The details of Neomero1 are presented elsewhere ^22^. The recruitment centers were in 6 different countries (Italy, Greece, Estonia, Spain, Lithuania, and UK), and local ethics committees provided approval. Informed consent was given by parents prior to enrollment and a separate consent was given for genetic studies for this work.

Detailed information is listed on EudraCT (2011-001515-31 and 2011-001521-25) and clinicaltrials.gov (NCT01551394 and NCT01554124).^21^ LOS was defined as sepsis in infants between 3 and 90 days of age.^21^ The diagnosis was established in one of two ways: 1) culture confirmed or 2) clinical sepsis.^2 3^ Culture confirmed cases had both an abnormal clinical or laboratory finding and a positive bacterial culture. Clinical Sepsis cases met clinical and laboratory criteria of Goldstein et al 2005^3^ or the 2010 EMA report^2^, but had a negative bacterial culture. All exclusion and inclusion criteria have been described previously^22^.

Controls were randomly selected from existing DNA banks in 3 of the European countries: Greece, Italy, and Spain from whom cases were recruited. Italian and Spanish controls were previously described adult population-based samples^23 24^. Greek controls were obtained from the Laboratory of Medical Genetics, Medical School, National & Kapodistrian, University of Athens, Greece, and were originally sent to the laboratory as part of a screening protocol for cystic fibrosis mutations. Although phenotypes were not available for the controls, these population-based controls should represent an appropriate comparator group as these individuals for the most part will not have had LOS due to its rarity and the likelihood of mortality for those who did contract it in these cohorts. Even at a prevalence of 4 per 1000^4^ only ∼1 control would be misclassified, causing a very small drop in power. All control samples were de-identified.

### Genotyping and Genomic Quality Control

DNA was extracted as previously described from 233 case blood samples (500 ul/sample)^25^. All samples were genotyped using the Illumina MEGA Consortium V2 Beadchip at the Hussman Institute for Human Genomics (University of Miami). Genome Studio V2011.1 (Illumina: San Diego, California) was used to generate the raw genotype calls. Standard quality control (QC) steps were performed^26-28^, and all of the following were removed prior to the initial association analyses: 1) SNPs missing calls in > 5% of participants, 2) participants with genotyping calls for less than 90% of the measured SNPs, 3) SNPs with minor allele frequencies < 5%, 4) participants with an excessively high or low number of heterozygous calls (> 3 SD away from the mean number of heterozygous calls for an individual) as individuals with a very low number of heterozygous calls may indicate inbreeding while a very high number may indicate sample contamination, and 5) participants who were cryptically related (pairs of samples with pihat > 0.2 were evaluated and one member of each pair was removed; this removed sample duplicates, full sibling, parent-child, and identical twin relationships from the data^29^. No pairs with pihat >0.2 remained after this step). It was important to eliminate relatives from the data because logistic regression analyses assume independent observations. After these steps there were 497 participants (224 cases, 273 controls) and 601876 autosomal/pseudo-autosomal SNPs available for association analyses (Figure S1). X chromosome SNPs not from the pseudo-autosomal region were handled separately as described below. After initial association analyses were performed, three additional QC checks were conducted on the top hits. SNPs were removed when there was evidence of: 1) deviation from Hardy-Weinberg Equilibrium (p < 1×10^−4^ among the controls), 2) differential patterns of missing SNPs between cases and controls (p <0.05 in a Fisher’s exact test), 3) evidence of SNP call errors in the raw data intensity plot from GenomeStudio (cluster separation score > 0.2).^28^

X chromosome SNPs were separated from the autosomal and pseudo-autosomal SNPs and QC steps described above were conducted separately in males and females. Subsequent association analyses were also conducted separately in males and females. 16664 SNPs passed the initial QC steps among the females (78 cases, 76 controls), and 15732 SNPs passed these steps among the males (146 cases, 197 controls). For the three post association QC steps: 1) the cluster separation scores were assessed as presented in the raw data (which included all participants), 2) differential patterns of missing SNPs between cases and controls was performed separately in the males and females, and 3) Hardy-Weinberg Equilibrium analyses were only performed for females.

### Statistical Analyses

We conducted logistic regression analyses on the 601876 autosomal/pseudo-autosomal SNPs and because none of the associations reached the canonical threshold for genome wide significance (p < 5×10^−8^) we present SNPs with p < 1×10^−4^ in each analysis. A similar approach has been utilized in prior GWAS of Neonatal Sepsis^30^, and may be required in GWAS of low incidence, severe/acute illness where the collection of large sample sizes is unlikely. Additionally, this method allows for hypothesis generation in the early stages of research, where type 2 error is most costly.^31^ Overall, the canonical p-value threshold (p < 5×10^−8^) has multiple limitations in genomic discovery settings ^32-36^ and we present results to facilitate further evaluation. We also performed logistic regression analyses on the top hits adjusting for sex and the first 4 principal components (to adjust for population substructure). We selected this number of principal components (PCs) because only the first 4 PCs had eigenvalues above 1.5 (Figure S2).

Neonatal sepsis is more common in males and infection susceptibility differs by sex^8-11^, thereby providing *a priori* reason to stratify analyses. This approach is critical from a biological perspective^8 10 14-16^ in the context of a trait with sex differences, but recent analytic guidelines strongly recommend considering the sexes independently, even when there is no *a priori* reason to suspect etiologic differences ^16-20^. Thus, we separated the males and females and reanalyzed the autosomal/pseudo-autosomal SNPs within sexes. Because top hits differed by sex, we evaluated a SNP-sex interaction term for each of the top hits (logistic regression model with terms for sex, the first 4 PCs, and SNP-sex interaction term) in the sex-combined data. This allowed us to determine if there was a significant difference in patterns of SNP-outcome association between the sexes.

Descriptive statistics were calculated using SAS (Cary, NC). QC and logistic regression analyses were conducted in Plink 1.07. PCs were obtained using Plink 1.9. Sex status was assigned with genetic confirmation using Plink 1.07. SNPs were assigned a gene annotation if they were intragenic or within 10Kb of a gene. Locus zoom (http://locuszoom.org/) was used to visualize regions around the top hits. Reactome (version 69, https://reactome.org) ^37 38^ was used to determine if the gene lists from our analyses were enriched for genes in known physiologic pathways.

### Validation of Prior Findings

Several published associations were reassessed. Srinivasan et al conducted a similar GWAS among extremely premature infants.^30^ The top ten SNPs from their work identified 4 regions of interest and we evaluated SNPs within 10 Kb of these loci. Additionally, a recent meta-analysis evaluated three candidate genes (*TNF*α, *IL6*, and *IL10*) ^39^, and we also evaluated SNPs that were intragenic or within 10Kb of these genes.

## Results

Altogether DNA samples from 233 patients with LOS and 288 controls were obtained for genetic analysis. After QC, 224 LOS cases and 273 adult controls remained (Figure S1); these included 197 (72.2%) male controls and 146 (65.2%) male cases (Table 1). Most LOS cases were born preterm (70.5%), 44.2% were culture confirmed, and the mean birth weight was 1.9 kg (Table 2).

**Table 1.** Characteristics of the 497 study participants.

|  | Study Population |  | Controls |  | Cases |  |
| --- | --- | --- | --- | --- | --- | --- |
|  | n | % | n | % | n | % |
| **Males** | 343 | 69.0 | 197 | 72.2 | 146 | 65.2 |
| **Females** | 154 | 31.0 | 76 | 27.8 | 78 | 34.8 |
| **Combined** | 497 | 100.0 | 273 | 54.9 | 224 | 45.1 |
| **Estonia** | 62 | 12.5 |  |  | 62 | 27.7 |
| **Greece** | 154 | 31.0 | 95 | 34.8 | 59 | 26.3 |
| **Italy** | 159 | 32.0 | 87 | 31.9 | 72 | 32.1 |
| **Lithuania** | 12 | 2.4 |  |  | 12 | 5.4 |
| **Spain** | 94 | 18.9 | 91 | 33.3 | 3 | 1.3 |
| **UK** | 16 | 3.2 |  |  | 16 | 7.1 |

**Table 2.**
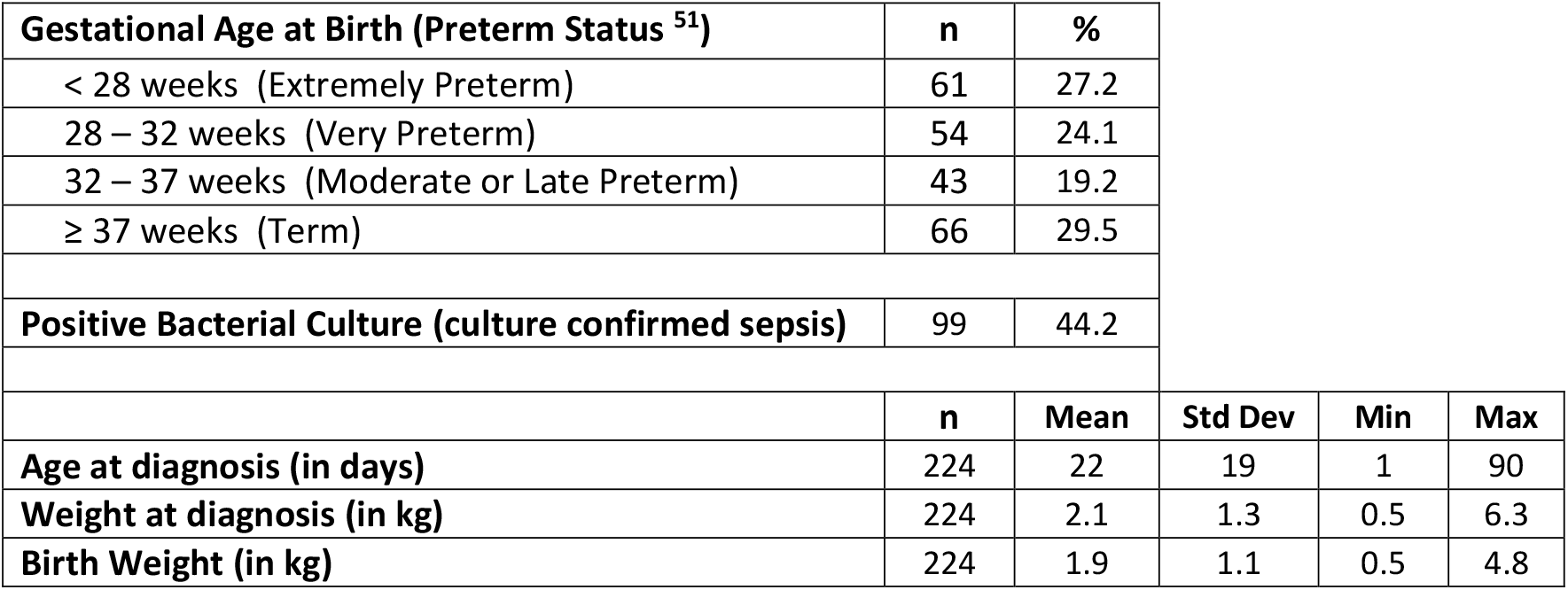
Characteristics of the neonatal sepsis cases.

| **Gestational Age at Birth (Preterm Status <sup>51</sup>)** | **n** | **%** |  |  |  |
| --- | --- | --- | --- | --- | --- |
| < 28 weeks (Extremely Preterm) | 61 | 27.2 |  |  |  |
| 28 – 32 weeks (Very Preterm) | 54 | 24.1 |  |  |  |
| 32 – 37 weeks (Moderate or Late Preterm) | 43 | 19.2 |  |  |  |
| ≥ 37 weeks (Term) | 66 | 29.5 |  |  |  |
| **Positive Bacterial Culture (culture confirmed sepsis)** | 99 | 44.2 |  |  |  |
|  | **n** | **Mean** | **Std Dev** | **Min** | **Max** |
| **Age at diagnosis (in days)** | 224 | 22 | 19 | 1 | 90 |
| **Weight at diagnosis (in kg)** | 224 | 2.1 | 1.3 | 0.5 | 6.3 |
| **Birth Weight (in kg)** | 224 | 1.9 | 1.1 | 0.5 | 4.8 |

### Analysis of male and female participants combined

In unadjusted logistic regression analyses, 37 SNPs from the autosomes and the pseudo-autosomal regions of the X chromosome were associated with LOS at p < 1×10^−4^ (Table 3), but none reached the canonical genome-wide significance level (p < 5×10^−8^). The p-values and odds ratios were similar after adjusting for sex and the first four principal components (Table 3). In all analyses the reference allele for OR calculations was the major allele. The 37 SNPs were annotated at the gene level (Table 3) and a pathway analysis was conducted using the curated Reactome Database^37 38^ (https://reactome.org/). A hypergeometric overrepresentation test indicated that this gene list was significantly enriched for components of 12 physiologic pathways (FDR <0.05); all these pathways involved NOTCH signaling (Table S1).

**Table 3:** Autosomal SNPs associated with neonatal sepsis among males and females combined ^a^.

| SNP and Chromosome |  | Logistic Regression <sup>b</sup> | Adjusted Logistic Regression <sup>c</sup> |  |  |  | Gene |
| --- | --- | --- | --- | --- | --- | --- | --- |
|  |  | p | OR | 95 % CI |  | p |  |
| rs7225568 | 17 | 2.83E-06 | 1.87 | 1.44 | 2.44 | 3.29E-06 | KCNH6 |
| rs7835808 | 8 | 3.21E-06 | 1.85 | 1.43 | 2.40 | 3.23E-06 | TRAPPC9 |
| rs597695 | 8 | 1.31E-05 | 0.57 | 0.44 | 0.74 | 1.99E-05 | Intergenic |
| rs16877997 | 6 | 1.61E-05 | 0.40 | 0.26 | 0.61 | 1.96E-05 | Intergenic locus chromosome 6 |
| rs12821366 | 12 | 1.91E-05 | 2.44 | 1.60 | 3.72 | 3.15E-05 | PIK3C2G |
| rs16877991 | 6 | 2.51E-05 | 0.42 | 0.28 | 0.63 | 3.40E-05 | Intergenic locus chromosome 6 |
| rs16832098 | 1 | 2.88E-05 | 0.29 | 0.16 | 0.51 | 2.44E-05 | BRINP3 |
| rs1386388033 | 17 | 2.97E-05 | 0.54 | 0.40 | 0.72 | 3.08E-05 | MAP2K4 <sup>d</sup> |
| rs72729202 | 1 | 3.72E-05 | 0.30 | 0.17 | 0.53 | 3.33E-05 | BRINP3 and LOC105371658 |
| rs10498688 | 6 | 4.24E-05 | 0.55 | 0.41 | 0.74 | 4.97E-05 | KIF13A |
| rs1568605 | 8 | 4.58E-05 | 0.59 | 0.46 | 0.76 | 4.60E-05 | TRAPPC9 |
| rs746456981 | 1 | 5.18E-05 | 0.31 | 0.17 | 0.54 | 4.41E-05 | BRINP3 |
| rs10877839 | 12 | 5.20E-05 | 0.57 | 0.43 | 0.75 | 7.68E-05 | Intergenic |
| rs13228161 | 7 | 5.24E-05 | 0.40 | 0.26 | 0.62 | 4.55E-05 | Intergenic |
| rs2076789 | 20 | 5.35E-05 | 0.55 | 0.41 | 0.73 | 5.87E-05 | Intergenic |
| rs4775425 | 15 | 5.35E-05 | 0.60 | 0.46 | 0.77 | 9.83E-05 | LOC107984782 |
| rs7845070 | 8 | 5.57E-05 | 1.69 | 1.31 | 2.19 | 6.47E-05 | TRAPPC9 |
| rs10743273 | 12 | 5.78E-05 | 1.74 | 1.33 | 2.29 | 6.53E-05 | PIK3C2G |
| rs28435105 | 15 | 6.02E-05 | 0.56 | 0.42 | 0.74 | 6.34E-05 | LOC101926994 <sup>d</sup> |
| rs1147855 | 6 | 6.12E-05 | 1.94 | 1.41 | 2.66 | 4.12E-05 | SASH1 |
| rs61737181 | 6 | 6.20E-05 | 0.41 | 0.26 | 0.64 | 7.51E-05 | HEY2 |
| rs78619534 | 4 | 6.41E-05 | 2.75 | 1.68 | 4.49 | 5.28E-05 | SCOC and SCOC-AS1 |
| rs7094526 | 10 | 6.43E-05 | 1.82 | 1.37 | 2.43 | 4.37E-05 | Intergenic |
| rs745172 | 6 | 6.82E-05 | 0.51 | 0.37 | 0.71 | 7.69E-05 | Intergenic locus chromosome 6 |
| rs1676959 | 8 | 7.05E-05 | 1.91 | 1.41 | 2.59 | 3.01E-05 | CSMD1 |
| rs146376406 | 12 | 7.28E-05 | 3.02 | 1.73 | 5.28 | 1.03E-04 | PIK3C2G |
| rs10900943 | 6 | 7.49E-05 | 1.88 | 1.38 | 2.55 | 5.50E-05 | Intergenic |
| rs13170530 | 5 | 7.56E-05 | 0.50 | 0.36 | 0.71 | 9.94E-05 | Intergenic |
| rs56168986 | 12 | 8.00E-05 | 2.29 | 1.52 | 3.47 | 8.57E-05 | Intergenic |
| rs11221264 | 11 | 8.40E-05 | 0.58 | 0.44 | 0.76 | 6.84E-05 | Intergenic |
| rs8118790 | 20 | 8.68E-05 | 2.08 | 1.44 | 3.00 | 9.05E-05 | SLC32A1 <sup>d</sup> |
| rs75362667 | 22 | 8.87E-05 | 0.43 | 0.28 | 0.66 | 1.16E-04 | ARFGAP3 <sup>d</sup> |
| rs16970766 | 17 | 8.93E-05 | 0.25 | 0.12 | 0.49 | 7.17E-05 | TNRC6C |
| rs1147859 | 6 | 8.93E-05 | 1.82 | 1.35 | 2.46 | 8.08E-05 | SASH1 |
| rs2414736 | 15 | 9.31E-05 | 1.68 | 1.29 | 2.18 | 1.21E-04 | LOC107984782 and LOC107984783 <sup>d</sup> |
| rs6539768 | 12 | 9.96E-05 | 2.30 | 1.51 | 3.51 | 9.85E-05 | Intergenic |
| rs11637386 | 15 | 9.98E-05 | 0.57 | 0.42 | 0.77 | 1.93E-04 | LOC107984782 |

<sup>a</sup> SNPs from autosomes and pseudo-autosomal regions of the x chromosome that associate at $p < 1 \times 10^{-4}$ <sup>b</sup> Allele dose coded logistic regression model<sup>c</sup> Allele dose coded logistic regression model adjusted for sex and the first 4 principal components<sup>d</sup> Intergenic but within 10Kb of this gene

### Analysis stratified by sex

In the sex-stratified logistic regression analyses 28 SNPs were detected in males (Table 4) and 16 SNPs in females (Table 5)(p < 1×10^−4^). Results remained similar after adjusting for the first four principal components. When evaluating the top male SNPs in females, none associated with LOS at p < 0.05 (Table 4), and none of the top female SNPs associated in males (Table 5). In combined data set analyses, 17 of the 28 male SNPs had a significant sex-SNP interaction term (p < 0.05; Table 4). All 16 of the top female SNPS had a significant sex-SNP interaction term (Table 5). The sex stratified analyses identified fewer SNPs and genes of interest at p < 1×10^−4^, and many of the loci identified in the sex-combined analysis were also top SNPs in the males-only analysis. No pathways were significantly overrepresented among the top hit genes for the males (FDR <0.05), and pathway analysis was uninformative among the females because only one of the genes is in the Reactome database. Top SNPs from the sex stratified analyses, and SNPs in LD with these top SNPs, had sex-dependent significance patterns (Figure S3-S6). Top SNPs from the sex-combined analysis, and SNPs in LD with these top SNPs, were more significant in the males-only analyses than in the females-only analyses (Figure S7-S8).

**Table 4:**
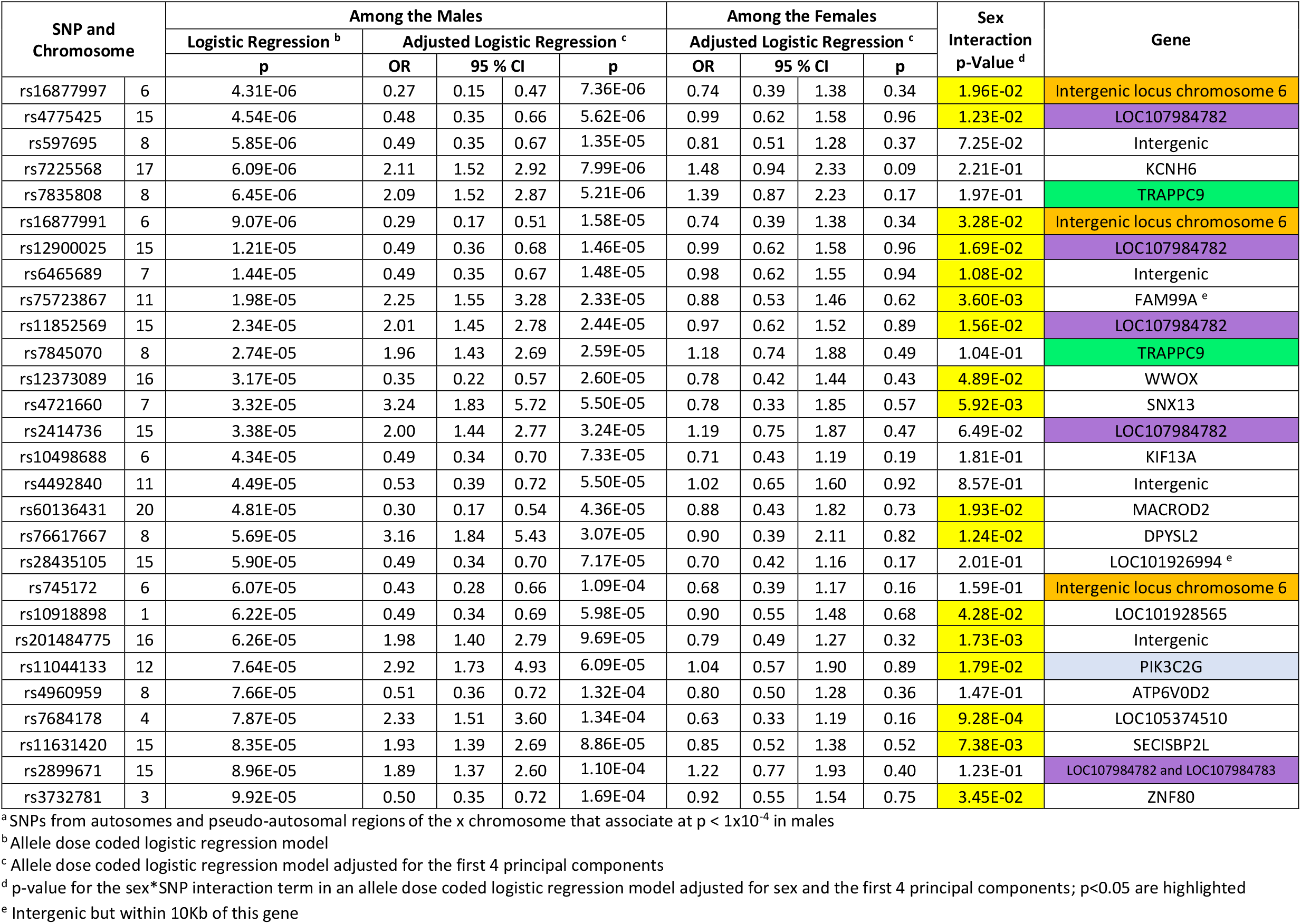
Autosomal SNPs associated with neonatal sepsis among the males ^a^.

**Table 5:**
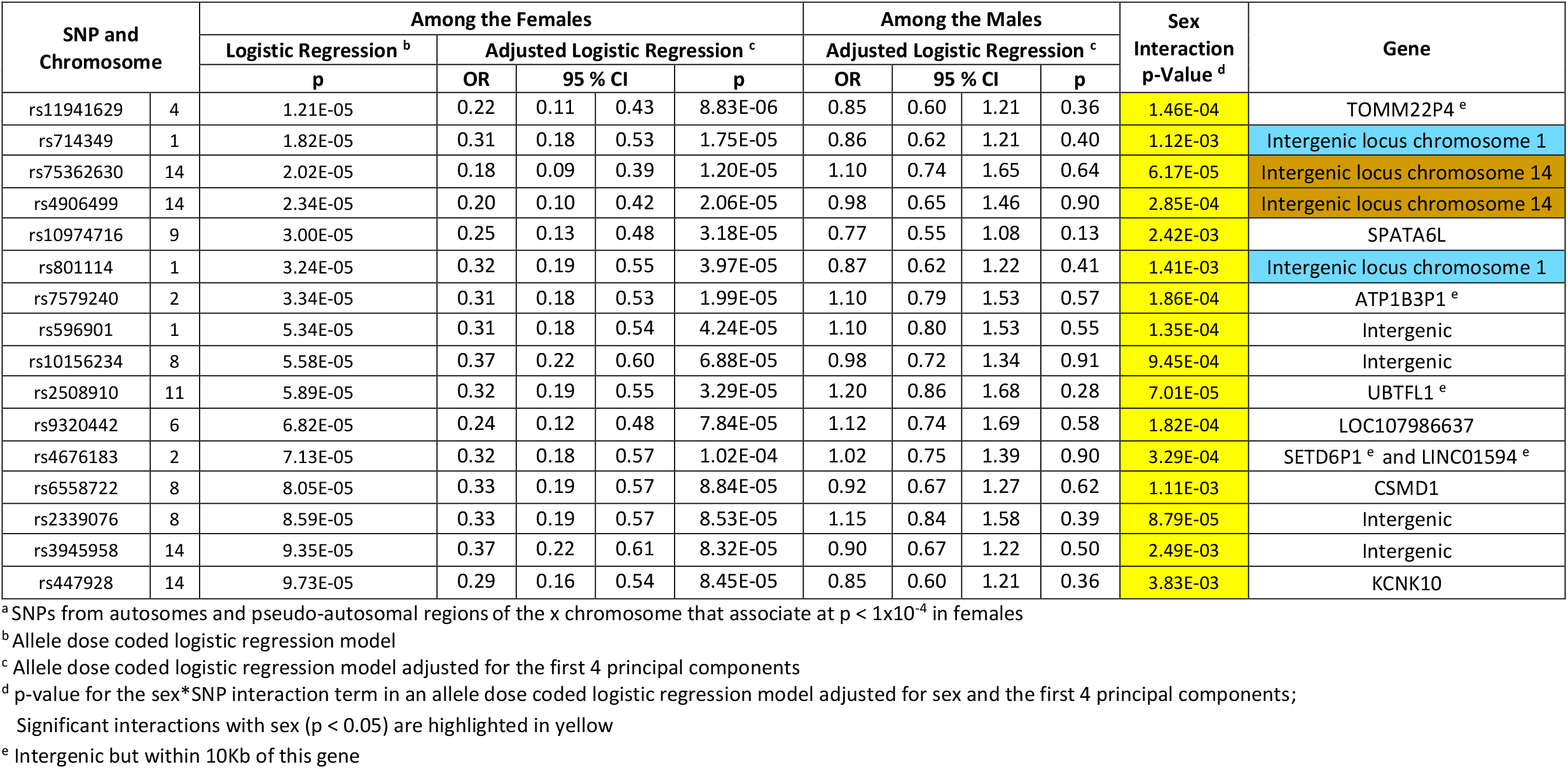
Autosomal SNPs associated with neonatal sepsis among the females ^a^.

### Analysis of X chromosome stratified by sex

No SNPs on the X chromosome significantly associated with LOS at p < 1×10^−4^. However, there were 13 SNPs in males and 9 SNPs in females that associated at p < 1×10^−3^. Similar to the pattern seen in the autosomal analysis, none of the top SNPs detected on the X chromosome in one sex replicated in the other sex (Table S2 and Table S3).

### Replication of Prior Findings

Four regions of interest on chromosome 2, 4, 6, and 16 were identified in a prior GWAS of neonatal sepsis.^30^ From these regions, we identified 3 SNPs that associated with neonatal sepsis (p < 0.05) on chromosome 16, and 1 SNP on chromosome 6 (Table 6). Three candidate genes of interest have been evaluated in prior meta-analyses.^39^ In evaluation of these three genes, we identified five SNPs from *IL10*, and one SNP from *TNF*α, that associated with LOS (Table 7, Figure S9). The five associating SNPs near *IL10* (intragenic +/-10kb) were in two linkage disequilibrium (LD) blocks (Figure S9). The minor alleles for rs1518111 and rs1800871, located near the transcription start site, were in strong LD (r^2^ = 0.9) and associated with decreased odds of neonatal sepsis (ORs 0.74 and 0.69, respectively). In contrast, the minor alleles for the 3’ SNPs, rs55705316, rs3024505, and rs3024493, were in LD (0.51 < r^2^ < 0.98) and were associated with increased odds of LOS (ORs 1.47 to 1.60).

**Table 6:**
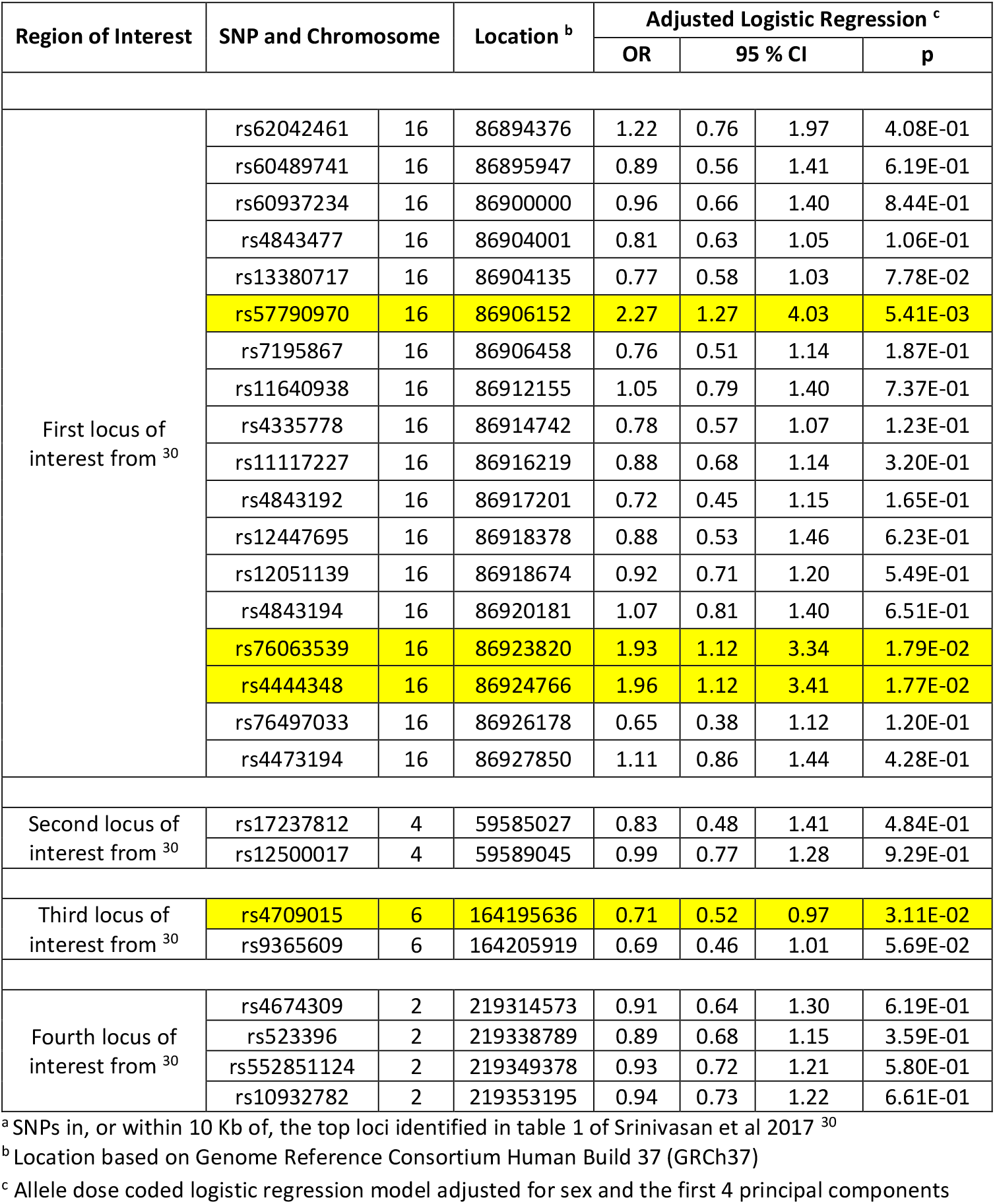
Reassessment of regions of interest from a prior GWAS ^a^.

**Table 7:** Reassessment of regions of interest from prior candidate gene studies ^a^.

| Region of Interest | SNP and Chromosome |  | Location <sup>b</sup> | Adjusted Logistic Regression <sup>c</sup> |  |  |  |
| --- | --- | --- | --- | --- | --- | --- | --- |
|  |  |  |  | OR | 95 % CI |  | P |
| Interleukin-10 <sup>39</sup> | rs13376708 | 1 | 206932503 | 0.79 | 0.57 | 1.10 | 1.65E-01 |
|  | rs76178457 | 1 | 206932693 | 0.73 | 0.45 | 1.18 | 2.00E-01 |
|  | rs55705316 | 1 | 206933517 | 1.47 | 1.01 | 2.14 | 4.70E-02 |
|  | rs111808242 | 1 | 206935478 | 1.16 | 0.90 | 1.50 | 2.62E-01 |
|  | rs61815632 | 1 | 206938439 | 1.07 | 0.77 | 1.49 | 6.95E-01 |
|  | rs3024505 | 1 | 206939904 | 1.60 | 1.10 | 2.31 | 1.33E-02 |
|  | rs3024493 | 1 | 206943968 | 1.58 | 1.09 | 2.30 | 1.71E-02 |
|  | rs1518111 | 1 | 206944645 | 0.74 | 0.55 | 1.00 | 4.80E-02 |
|  | rs1800871 | 1 | 206946634 | 0.69 | 0.52 | 0.93 | 1.43E-02 |
|  | rs1800896 | 1 | 206946897 | 1.17 | 0.90 | 1.53 | 2.39E-01 |
| Tumor Necrosis Factor <sup>39</sup> | rs2857602 | 6 | 31533378 | 1.02 | 0.79 | 1.32 | 8.72E-01 |
|  | rs2844484 | 6 | 31536224 | 1.02 | 0.79 | 1.32 | 8.72E-01 |
|  | rs2009658 | 6 | 31538244 | 0.97 | 0.69 | 1.38 | 8.69E-01 |
|  | rs915654 | 6 | 31538497 | 0.89 | 0.68 | 1.17 | 4.13E-01 |
|  | rs1800683 | 6 | 31540071 | 0.82 | 0.61 | 1.11 | 2.01E-01 |
|  | rs909253 | 6 | 31540313 | 0.83 | 0.62 | 1.11 | 2.10E-01 |
|  | rs2229094 | 6 | 31540556 | 1.16 | 0.87 | 1.53 | 3.09E-01 |
|  | rs2229092 | 6 | 31540757 | 1.03 | 0.61 | 1.75 | 9.02E-01 |
|  | rs1041981 | 6 | 31540784 | 0.81 | 0.60 | 1.09 | 1.65E-01 |
|  | rs1041981 | 6 | 31540784 | 0.78 | 0.58 | 1.06 | 1.10E-01 |
|  | rs1799964 | 6 | 31542308 | 1.23 | 0.90 | 1.68 | 1.85E-01 |
|  | rs1799964 | 6 | 31542308 | 1.23 | 0.90 | 1.68 | 1.85E-01 |
|  | rs1800630 | 6 | 31542476 | 0.93 | 0.66 | 1.32 | 6.83E-01 |
|  | rs1799724 | 6 | 31542482 | 0.94 | 0.66 | 1.33 | 7.22E-01 |
|  | rs1800629 | 6 | 31543031 | 0.75 | 0.50 | 1.12 | 1.62E-01 |
|  | rs361525 | 6 | 31543101 | 2.20 | 1.26 | 3.83 | 5.74E-03 |
|  | rs1800610 | 6 | 31543827 | 0.95 | 0.68 | 1.34 | 7.74E-01 |
|  | rs3093662 | 6 | 31544189 | 1.53 | 0.99 | 2.36 | 5.44E-02 |
|  | rs3093664 | 6 | 31544642 | 1.32 | 0.82 | 2.11 | 2.48E-01 |
|  | rs769178 | 6 | 31547514 | 1.08 | 0.75 | 1.55 | 6.71E-01 |
|  | rs769177 | 6 | 31547611 | 0.90 | 0.52 | 1.57 | 7.05E-01 |
| Interleukin-6 <sup>39</sup> | rs6461662 | 7 | 22757089 | 0.90 | 0.69 | 1.17 | 4.24E-01 |
|  | rs13438415 | 7 | 22760002 | 1.06 | 0.75 | 1.50 | 7.49E-01 |
|  | rs10499563 | 7 | 22760488 | 1.04 | 0.77 | 1.41 | 8.09E-01 |
|  | rs1800797 | 7 | 22766221 | 0.95 | 0.72 | 1.26 | 7.38E-01 |
|  | rs1800796 | 7 | 22766246 | 0.69 | 0.42 | 1.12 | 1.35E-01 |
|  | rs1800795 | 7 | 22766645 | 0.96 | 0.72 | 1.28 | 7.79E-01 |
|  | rs2069861 | 7 | 22771654 | 0.94 | 0.55 | 1.62 | 8.21E-01 |

<sup>a</sup> SNPs in, or within 10 Kb of, the candidate genes evaluated in Srinivasan et al 2017 <sup>39</sup><sup>b</sup> Location based on Genome Reference Consortium Human Build 37 (GRCh37)<sup>c</sup> Allele dose coded logistic regression model adjusted for sex and the first 4 principal components

## Discussion

We identified 71 SNPs that associated with LOS at p < 1×10^−4^ in at least one of five regression analyses: 1) sex-combined-autosomal, 2) males-only-autosomal, 3) females-only-autosomal, 4) males-only-X-chromosomal, and 5) females-only-X-chromosomal. However, none were significant at the canonical genome-wide significance level. Overall, this was not unexpected as the sample size was modest, the outcome was complex, and exposure to infectious agents varied. Nonetheless, many biologically meaningful association signals will not reach this canonical threshold ^40 41^ and requiring this level of significance can be problematic for discovery.^31 36^ Thus, the top hits in these analyses may still be of interest if they are corroborated with diverse convergent evidence.^36^ In other words, statistical evidence from high dimensional observational epidemiology (e.g. GWAS based associations) is of greater interest when informatic evidence (e.g. pathway analyses in curated biological databases) and laboratory experimentation (e.g. transgenic knockout models or in vitro biochemical experiments) can validate the findings. Here we use the DiCE metric^36^ to characterize the diversity and strength of validation for the biological signals. Overall, we observed three key patterns that have implications for further research: 1) links between LOS and *IL10* and two uncharacterized genes on chromosomes 16 and 6; 2) the pattern of genes identified in the sex-combined-autosomes analysis implicates NOTCH signaling; and 3) many of the observed associations differed compellingly by sex.

### Comparisons to prior genetic analyses of neonatal sepsis

Although our analyses did not reveal genome-wide significant associations, they were still useful for replicating prior findings. However, the existing studies evaluated males and females together, and this approach should be expected to detect variants that modify susceptibility in both sexes in the same direction, but not variants that modify susceptibility only in one sex. Hence, we compared the existing literature to only the findings from our sex-combined analysis because our results indicate that a precision medicine approach to LOS will necessitate stratifying by sex.

Most relevant literature in LOS comes from candidate gene studies, and three of the most studied genes (*TNF*α, *IL6*, and *IL10*) were recently evaluated in a meta-analysis. Evidence indicated trends for *TNF*α and *IL10*, but only *IL10* reached significance.^39^ We identified a similar pattern in our combined male/female analysis: no *IL6* SNPs associated with neonatal sepsis but 5 *IL10* SNPs, and 1 *TNF*α SNP associated at p <0.05 (Table 7). This convergence with prior meta-analyses serves to validate both our approach and novel findings. Importantly, we identified two LD blocks among the five significant SNPs in the *IL10* region. The minor alleles for rs1518111 and rs1800871 are located near the transcription start site and were associated with decreased odds of neonatal sepsis, while the minor alleles for the three SNPs near the 3’ end of gene (rs55705316, rs3024505, and rs3024493) associated with increased odds of neonatal sepsis. These findings reinforce the putative importance of *IL10* in LOS and they may be used to develop testable molecular hypotheses for animal or in vitro models.

In addition to prior work with candidate genes, at least one GWAS has been conducted on neonatal sepsis.^30^ This study was restricted to extremely premature infants and it did not include sex stratified analyses, so the comparisons to our study are limited. Yet, we found corroborating evidence in our sex-combined analysis for 2 of the 4 loci identified (Table 6). In short, we identified 3 significant SNPs (p<0.05) in LOC105371393 on chromosome 16, overlapping with the prior study. Given the variation in the phenotypes between our and the previous GWAS, the results indicate that the genes in common are likely to be strong modulators of sepsis risk operating across sub-phenotypes. While this locus remains uncharacterized molecularly, the association with neonatal sepsis in an independent GWAS is intriguing. Thus, this locus currently has a DiCE score of 3 based on omic/observational evidence alone.^36^ Because no informatic or laboratory data yet exists for this locus, 4 is the maximum possible score at this stage in the research process; thus, additional research on this region is warranted. We also identified a significant SNP (p<0.05) in LOC107986666 of chromosome 6. Again, this uncharacterized locus has a DiCE score of 3 in a setting where a maximum of 4 points are possible, thus further investigation of this region is also warranted. Details of the DiCE metric calculation are available^36^, but in brief, the current DiCE score of 3 was achieved as follows: 3 points for statistical associations observed in two separate GWAS, 0 points for informatic evidence, and 0 points for experimental evidence.

### NOTCH signaling in Neonatal Sepsis

In pathway analyses the gene list from the sex-combined autosome analysis was significantly enriched for components of 12 physiologic pathways all of which implicated NOTCH signaling (Table S1). Overall, this supports further research into the role of NOTCH signaling in sepsis, as well as, the existing small-molecule modulators of NOTCH signaling.^42-49^

### Sex Differences in Neonatal Sepsis

None of the top SNPs in males were associated with sepsis in the females (at p<0.05), and none of the top SNPs in females associated with sepsis in males (at p<0.05). These statements hold for both the autosomal and X chromosomal variants. This pattern is quite striking and was corroborated with formal statistical interaction analyses in the sex-combined-autosomes data. Seventeen of the 28 hits among the males, and 16 of the 16 hits among the females, significantly interacted with sex. Furthermore, all 16 of the sex*SNP interaction terms among the females were highly significant (p<1×10^−3^). This pattern of interactions is quite remarkable given the modest size of our dataset; the probability of having all these interaction terms be significant is of the order of 10^−48^. Additionally, the locus zoom plots reveal that the SNPs in LD with the top hit SNPs from the stratified analyses have sex dependent significance patterns, corroborating patterns observed for the top hits. We note that we could not combine males and females to evaluate interaction terms for the X-chromosome SNPs as such analyses are biologically and analytically incompatible. Lastly, we found that associations among the males had excess influence in driving the results of the sex-combined analysis. Specifically, we observed that the identified genes of interest, and the SNP significance patterns were similar in the sex-combined and males-only analyses (Figure S5-S8). This finding is not surprising as males are more susceptible to sepsis^8-11^ and there were more males in our study. However, males may have also been overrepresented in prior studies of neonatal sepsis that did not stratify by sex.

Overall, the prominent sex differences we observed indicate that some components of the etiology of LOS are different in males and females, and future research in this area should always consider the sexes separately. The existence of sex specific pathophysiologies in neonatal sepsis ^8-11^ was proposed over 50 years ago, but this is often ignored in neonatal sepsis research. Failing to attend to this may have hampered prior studies due to etiological heterogeneity. However, the sex of the neonates is nearly always recorded, and thus this heterogeneity is retrospectively addressable. In summary, our findings suggest that sex specific analyses should be conducted routinely in neonatal sepsis research.

### Weaknesses and Strengths

Several study weaknesses and strengths deserve mention. One weakness was that we were not able to obtain controls from neonates in the six European countries, but we obtained adult controls from three of the six countries (Greece, Italy, and Spain). However, this control selection strategy is not likely to generate meaningful misclassification or bias because the incidence of neonatal sepsis in high income countries is 2-4 per 1000 live births.^4^ Thus, we would expect that no more than 1 of the 273 controls to be misclassified (273 × 0.004 = 1.092). A second weakness of this study is its small sample size (497 participants and 224 cases) and we would expect this to reduce power to both replicate prior findings and to find novel associations. However, despite limited power, we have identified consistent, significant patterns that corroborate and extend prior findings. A strength and weakness is that all of our samples were from Europe. The relatively homogenous study populations did not show substantial ancestry variation and this should increase the power and validity of our analyses among Europeans but may reduce generalizability to other ancestry groups^50^. Finally, we emphasize that our sex stratification approach allowed us to address a critical but underappreciated source of etiologic heterogeneity that has hindered neonatal sepsis research.

## Conclusions

We studied the role of genetics in LOS using a GWAS strategy and made several key observations. First, our sex-combined association analysis identified 37 SNPs of interest and corroborated previously identified links between neonatal sepsis and *IL10* as well for two uncharacterized loci from chromosomes 6 and 16. Second, our pathway analyses motivates exploration of interventions that utilize small molecule modulators of NOTCH signaling. Third, our sex stratified analyses revealed 28 SNPs of interest among only males and 16 non-overlapping SNPs of interest among only females. Most of the top male SNPs and all the top female SNPs demonstrated statistically significant interactions with sex. Overall, these findings add to the growing evidence that male and female infants differ in their physiologic response to infectious disease, and they should routinely be considered separately in future studies of neonatal sepsis.

## Supporting information

Strobe Checklist - Neonatal Sepsis GWAS

## Data Availability

Data is not generally available due to ethical considerations but may be requested from the authors.

## NeoMero Consortium

Jean Pierre Aboulker, Oguz Akbas, Antonella Allegro, Cinzia Auriti, Chiara Bertaina, Davide Bilardi, Giulia Bonatti, Fuat Emre Canpolat, Francesca Ippolita Calo Carducci, Corine Chazallon, Nijole Drazdienė, Susanna Esposito, Silvia Faggion, Isabelle Fournier, Eva Germovsek, Carlo Giaquinto, Genny Gottardi, Hayriye Gözde, Tiziana Grossele, Maarja Hallik, Cristina Haass, Paul Heath, Tatiana Munera Huertas, Valentina Ierardi, Mari-Liis Ilmoja, Elias Iosifidis, Sandrine Kahi, Paraskevi Karagianni, Aspasia Katragkou, Eve Kaur, Birgit Kiilaspää, Karin Kipper, Aggeliki Kontou, Victoria Kougia, Kanmaz Kutman, Jelena Kuznetsova, Elisabetta Lolli, Tuuli Metsvaht, Laurence Meyer, George Mitsiakos, Valentina Montinaro, Fabio Mosca, Makis Mylonas, Kader Ben Abdelkader Emmanuelle Netzer, Clarissa Oeser, Felix Omenaca, Zoi Dorothea Pana, Maria Luisa Paoloni, Simona Perniciaro, Laura Picault, Carlo Pietrasanta, Lorenza Pugni, Andrea Ronchi, Paolo Rossi, Suzan Şahin, Yacine Saidi, Laura Sanchez, Kosmas Sarafidis, Michael Sharland, Marina Spinelli, Joseph Standing, Claudia Tagliabue, Tuuli Tammekunn, Nina Tiburzi, Ursula Trafojer, Vytautas Usonis, Adilia Warris.

## Supplemental Materials

**Figure S1:**
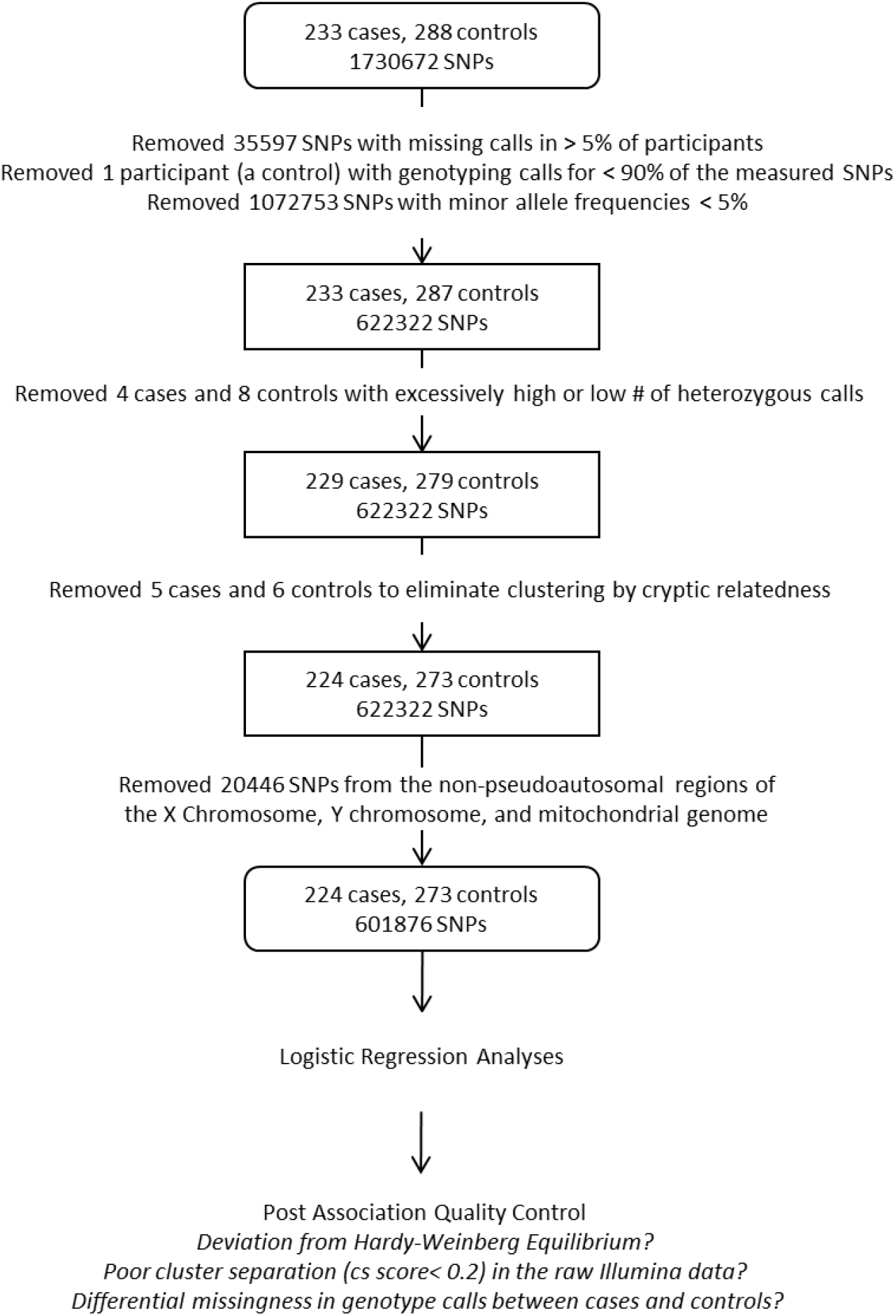
Quality Control Procedures Flow Chart.

**Figure S2:**
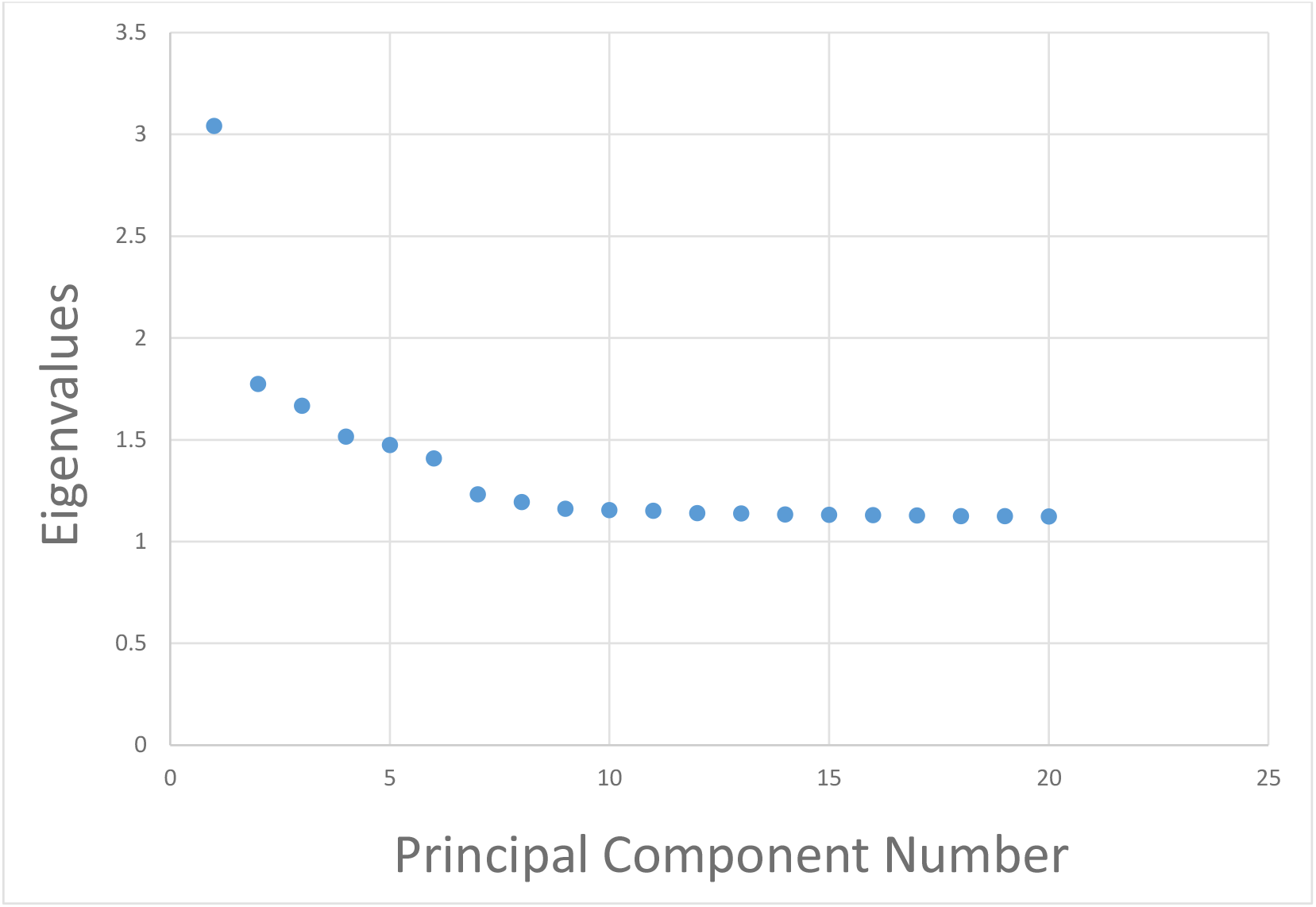
Scree Plot of Eigenvalues for the Principal Components.

**Table S1:** Reactome based pathway analysis for genes identified in the combined sex autosome analysis (Table 1) 12 physiologic pathways are better represented in the gene list than would be expected by chance (FDR <0.05). All of these pathways involve NOTCH signaling. Note that BRINP3, CSMD1, SASH1 were not in the curated Reactome database and this analysis was therefore based on the remaining gene hits: KCNH6, TRAPPC9, PIK3C2G, MAP2K4, KIF13A, HEY2, SCOC, SLC32A1, ARFGAP3, TNRC6C. https://reactome.org/PathwayBrowser/#TOOL=AT

| Pathway name | Hypergeometric Test for overrepresentation of the pathway |  |
| --- | --- | --- |
|  | p-Value | FDR <sup>a</sup> |
| NOTCH4 Intracellular Domain Regulates Transcription | 3.02E-04 | 0.030 |
| NOTCH3 Intracellular Domain Regulates Transcription | 5.75E-04 | 0.030 |
| NOTCH1 Intracellular Domain Regulates Transcription | 1.38E-03 | 0.030 |
| Signaling by NOTCH3 | 1.74E-03 | 0.030 |
| Signaling by NOTCH | 1.87E-03 | 0.030 |
| Signaling by NOTCH1 HD+PEST Domain Mutants in Cancer | 2.02E-03 | 0.030 |
| Constitutive Signaling by NOTCH1 HD+PEST Domain Mutants | 2.02E-03 | 0.030 |
| Constitutive Signaling by NOTCH1 PEST Domain Mutants | 2.02E-03 | 0.030 |
| Signaling by NOTCH1 PEST Domain Mutants in Cancer | 2.02E-03 | 0.030 |
| Signaling by NOTCH1 in Cancer | 2.02E-03 | 0.030 |
| Signaling by NOTCH1 | 2.98E-03 | 0.039 |
| Signaling by NOTCH4 | 3.64E-03 | 0.044 |

<sup>a</sup> probability score corrected for false discovery rate by the Benjamani-Hochberg method

**Figure S3.**
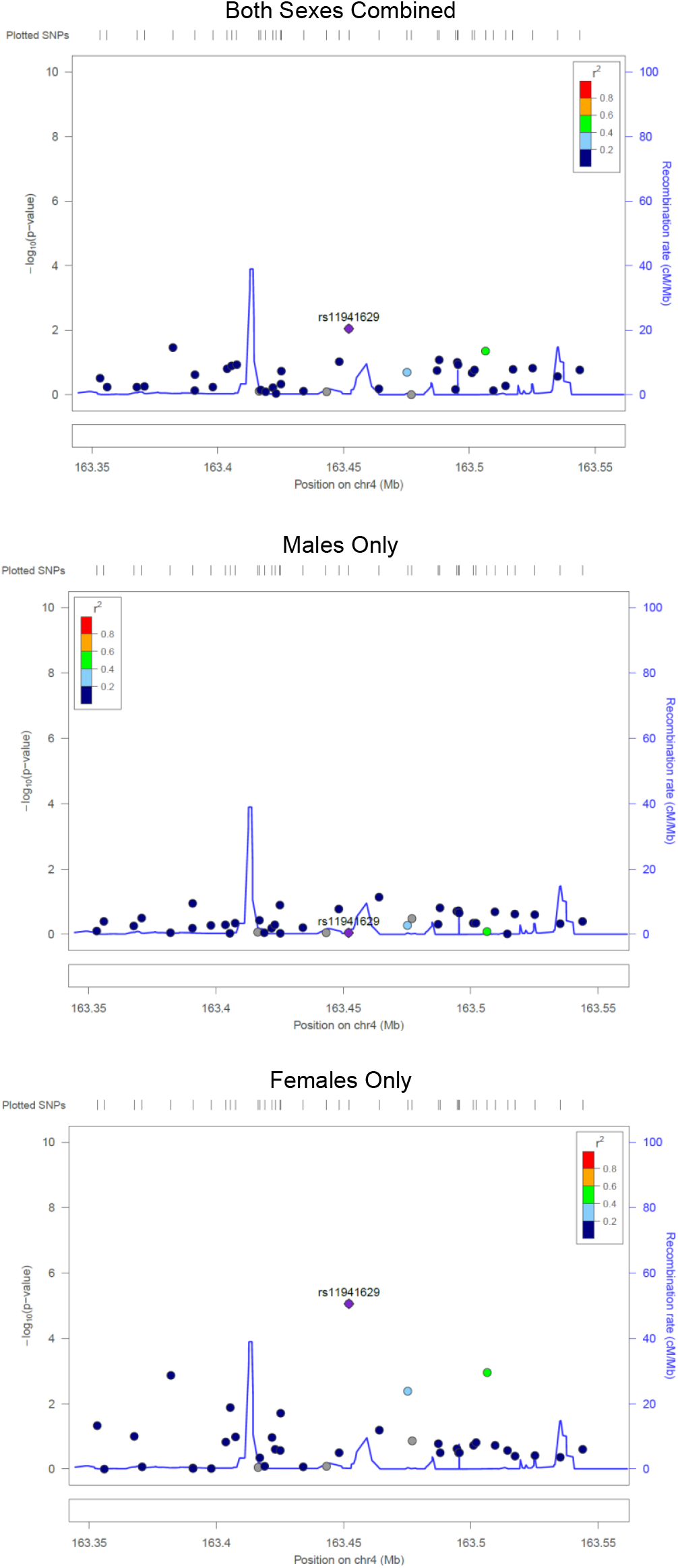
Locus Zoom Plots displaying p-values and LD structure around the top hit in the females-only analysis.

**Figure S4.**
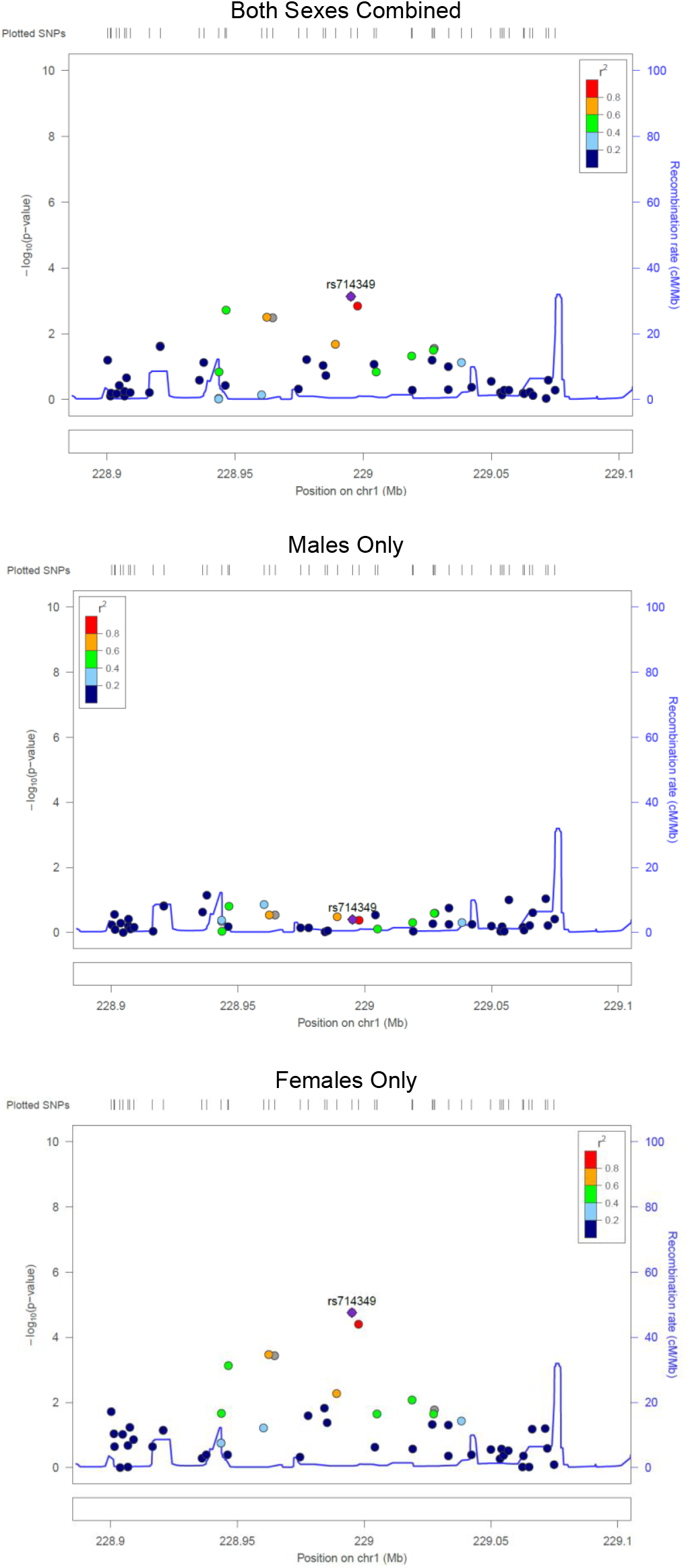
Locus Zoom Plots displaying p-values and LD structure around the second hit in the females-only analysis.

**Figure S5.**
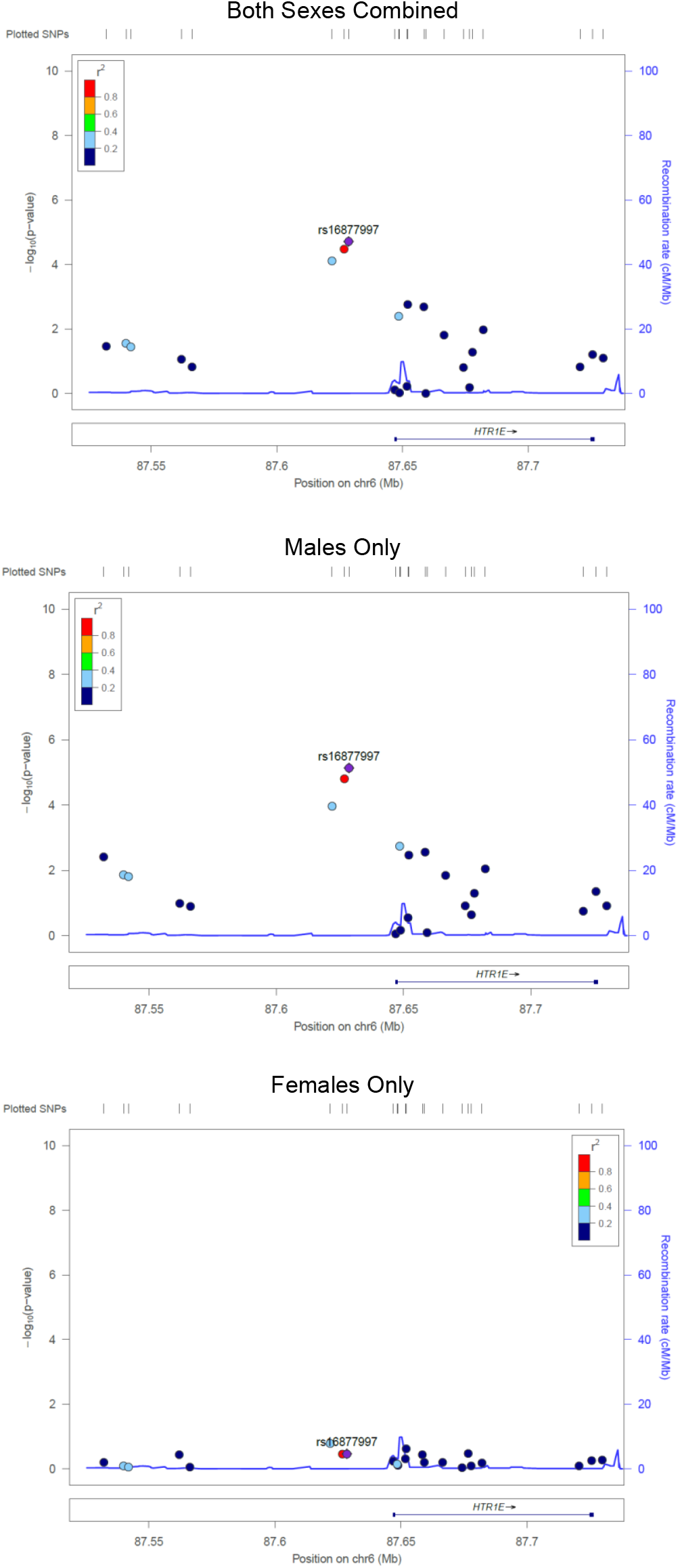
Locus Zoom Plots displaying p-values and LD structure around the top hit in the males-only analysis.

**Figure S6.**
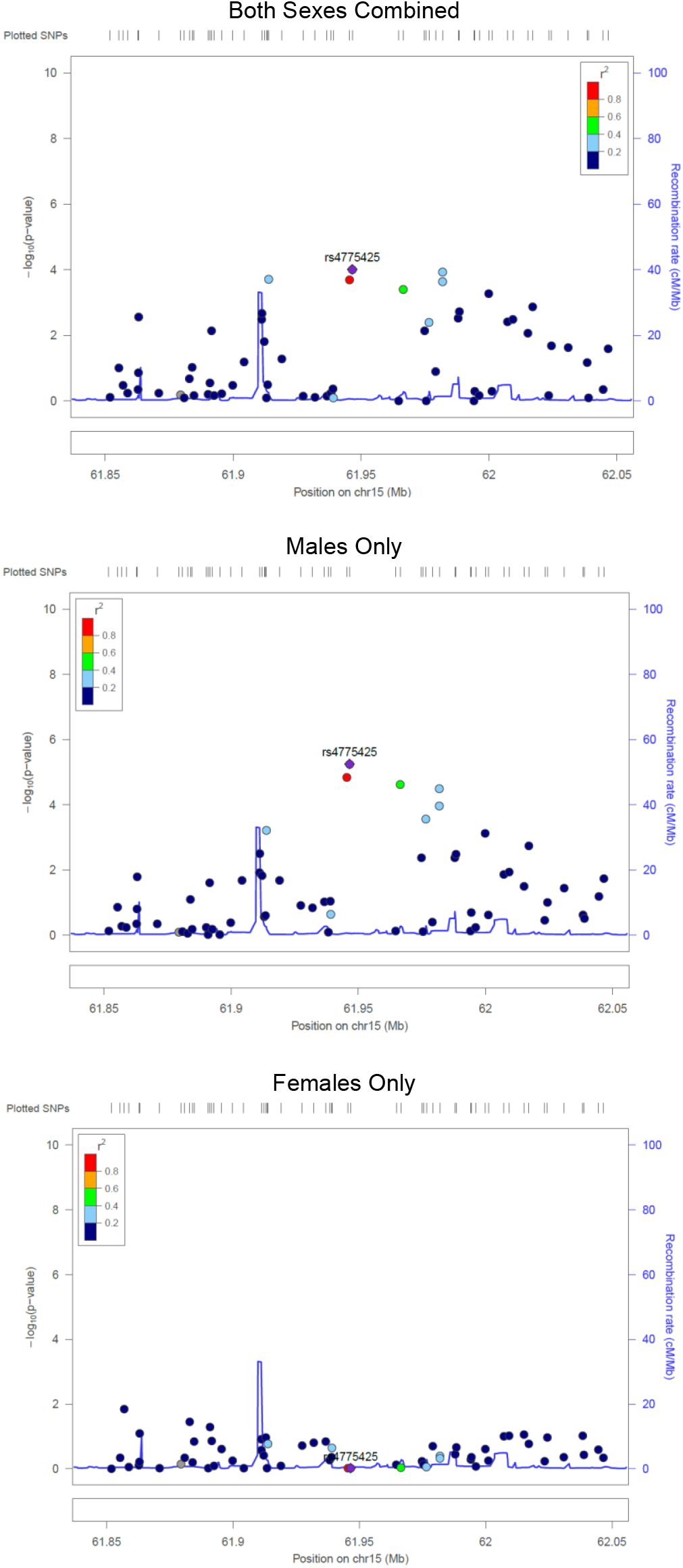
Locus Zoom Plots displaying p-values and LD structure around the second hit in the males-only analysis.

**Figure S7.**
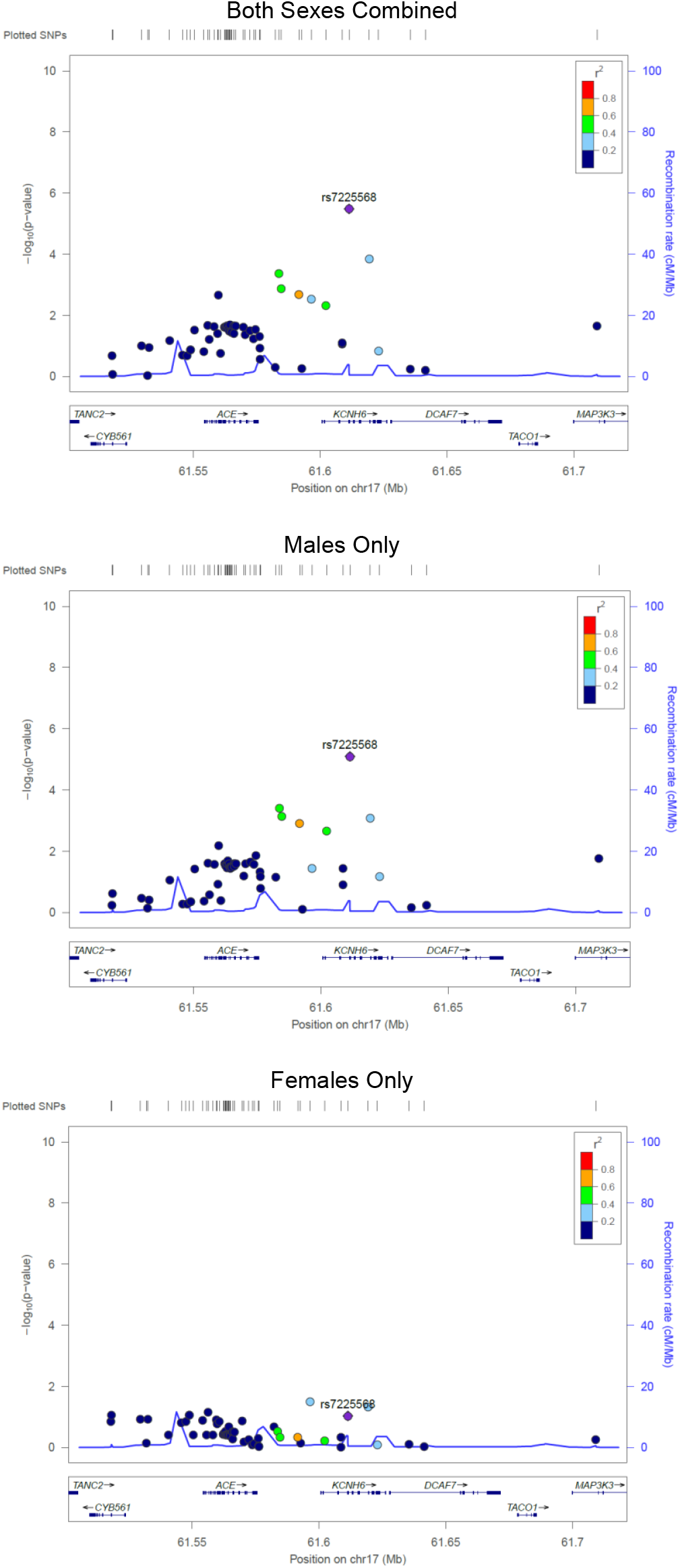
Locus Zoom Plots displaying p-values and LD structure around the top hit in the sex-combined analysis.

**Figure S8.**
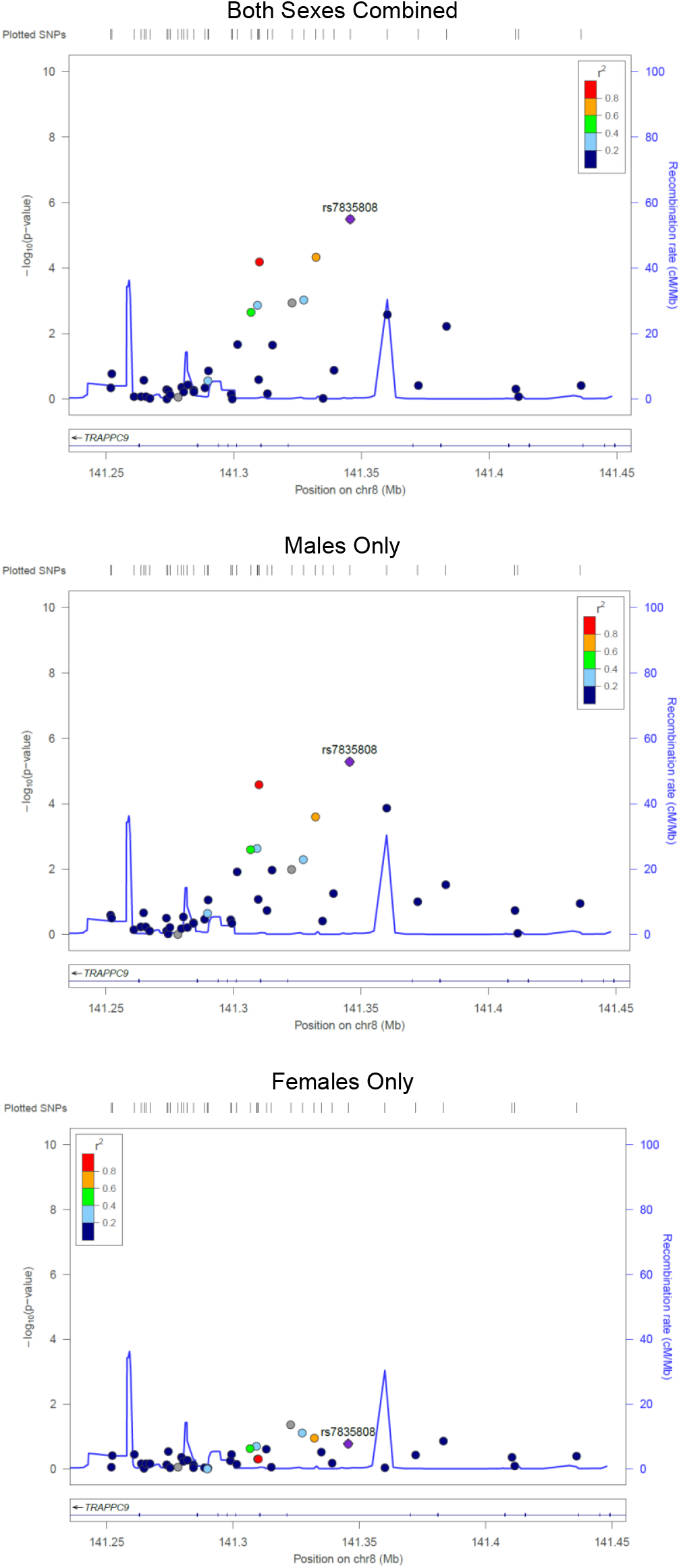
Locus Zoom Plots displaying p-values and LD structure around the second hit in the sex-combined analysis.

**Table S2:**
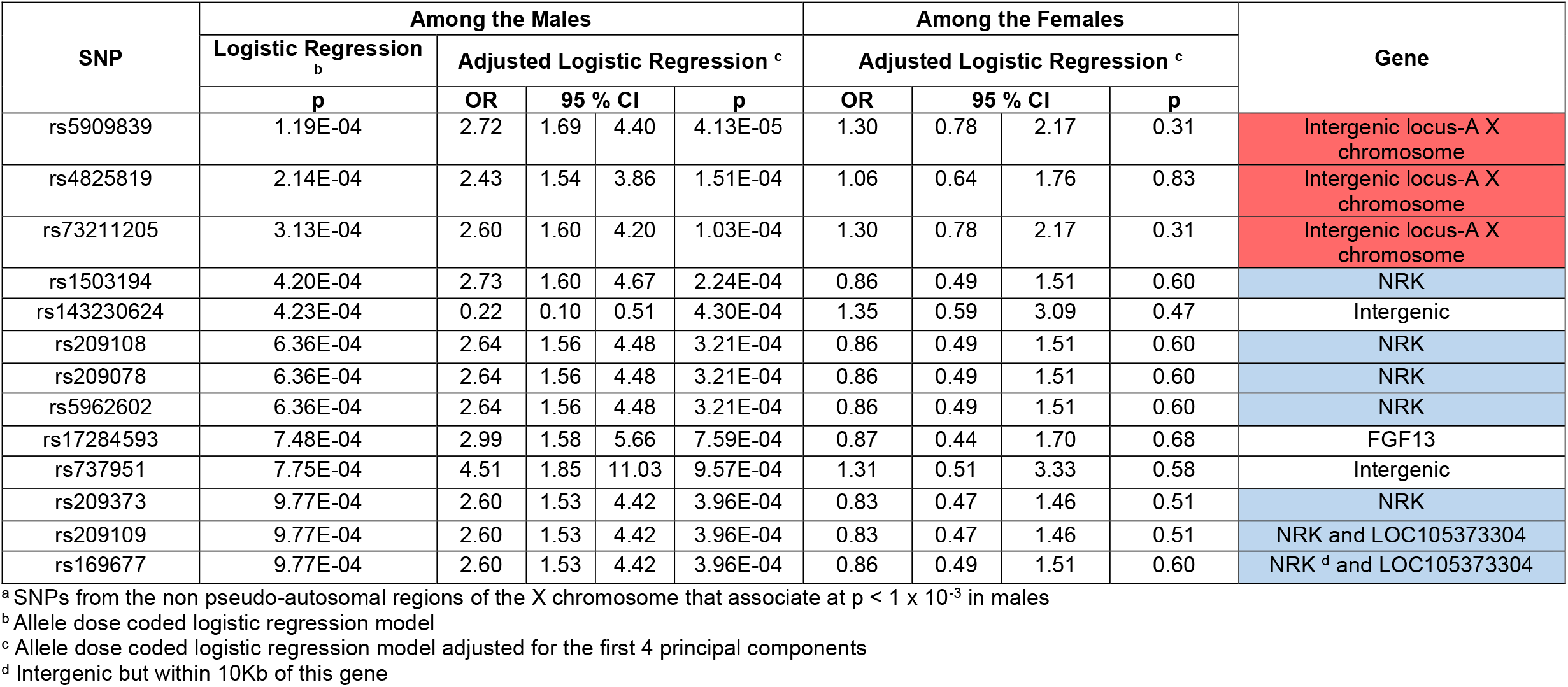
X chromosome SNPs associated with neonatal sepsis among the males ^a^.

**Table S3:**
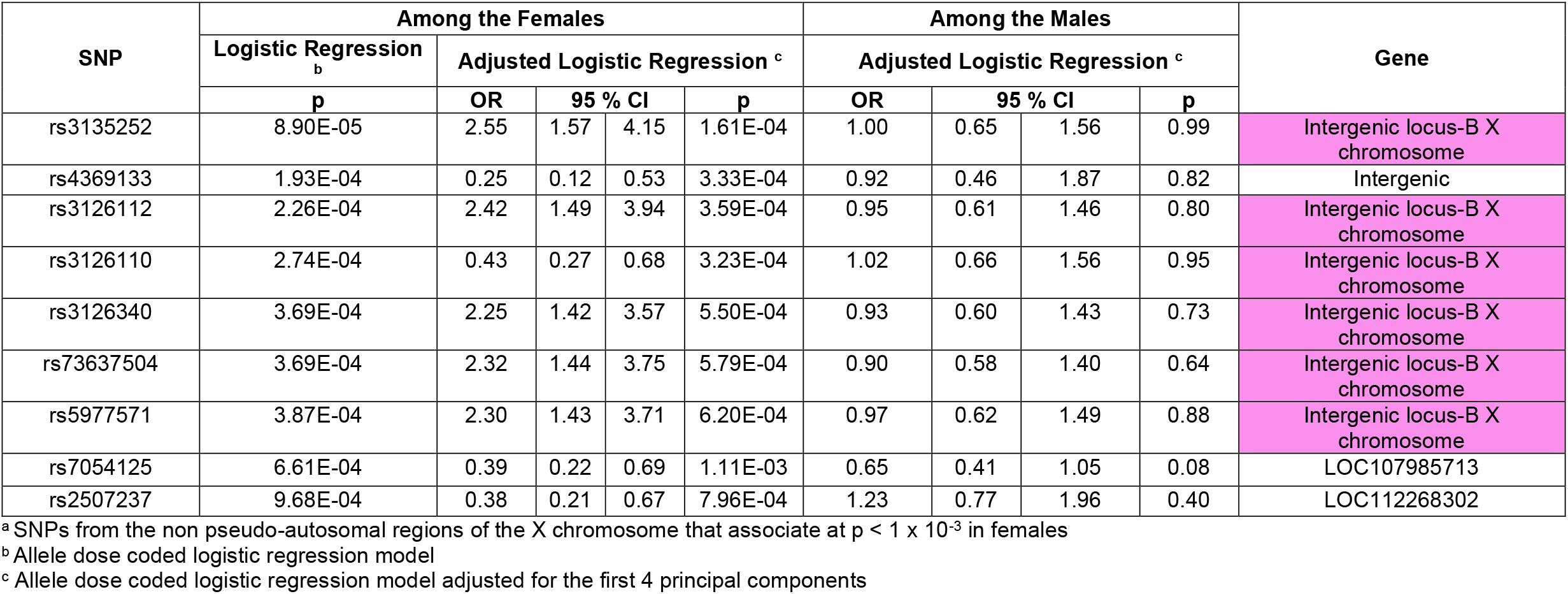
X chromosome SNPs associated with neonatal sepsis among the females ^a^.

**Figure S9:**
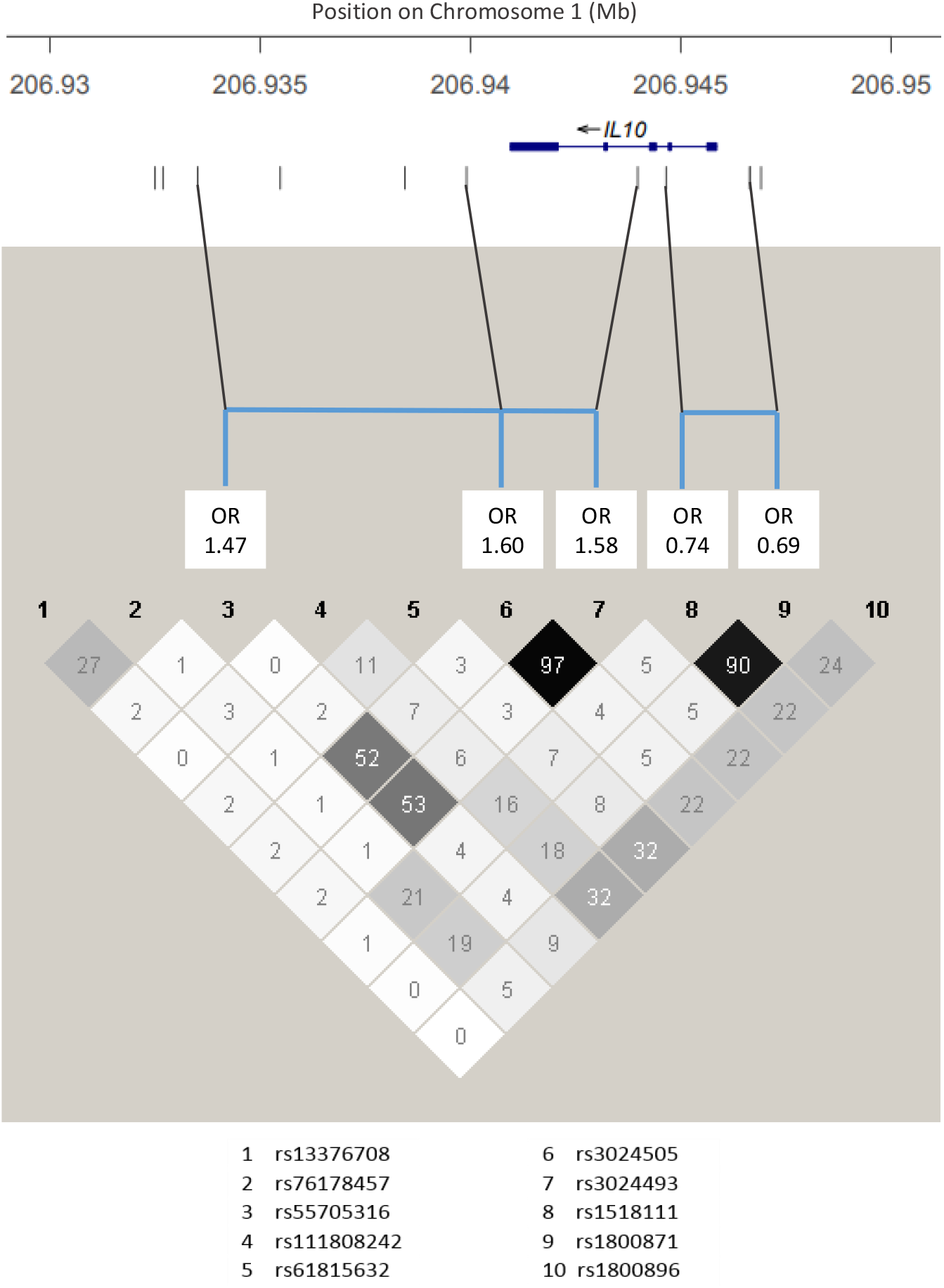
Haploview plot of the LD structure for the IL-10 SNPs with the relative genomic location overlaid from the Locus Zoom Plot. LD structure is displayed with r^2^ values and ORs for SNPs with significant associations in the sex-combined autosomal analysis (p<0.05) are presented above their location on the plot. In all analyses the reference allele for OR calculations is the major allele. Thus, rs55705316, rs3024505, and rs3024493 are in LD, and their minor alleles are associated with increased odds of neonatal sepsis. Additionally, rs1518111 and rs1800871 are in LD, and their minor alleles are associated with decreased odds of neonatal sepsis.

